# Decline to near-zero malaria hospitalisation over 35 years on the Kenyan Coast

**DOI:** 10.64898/2026.07.13.26357922

**Authors:** Alice Kamau, Moses M Musau, Neema Mturi, Shebe Mohammed, Gabriel Mwambingu, David Walumbe, Amek Nyaguara, Thomas N Williams, Philip Bejon, Robert W Snow

## Abstract

**Background:** Few longitudinal studies have examined the long-term health impact of changing malaria prevention and treatment strategies in Africa. This study analyses 35 years of clinical surveillance among paediatric malaria admissions in Kilifi, Kenya.

**Methods:** Children aged 1 month to 14 years who were residents of rural Kilifi Health and Demographic Surveillance System and admitted to Kilifi County Hospital between January 1990 and December 2024 were included. Community malaria exposure was estimated using infection prevalence among children admitted for trauma, elective surgery, bites and neoplasms. Malaria admissions were defined as hospitalisations with a positive blood slide and a primary, secondary, or co-morbid malaria diagnosis. Severe malaria phenotypes including severe anaemia, cerebral malaria, hyper-parasitaemia and in-hospital mortality were also examined. Binomial and Poisson regression models assessed temporal differences, using 1990–1996 as the reference period.

**Results:** Community malaria prevalence declined from 35% (95% CI: 31, 39) in the 1990s to 2% (95% CI: 1, 4; p<0.001) in 2020–2024. Malaria hospitalisations declined from a peak of 25.5 per 1,000 children per annum (p.a.) (95% CI: 24.4–26.6) in 1999 to 0.65 (95% CI: 0.59–0.72; p<0.001) between 2020–2024. The median age of malaria increased from 19 months (IQR: 12, 39) between 1990-1996 to a peak of 48 months (IQR: 28, 78) between 2012-2019. Cerebral malaria became proportionally more common than severe anaemia over time. Malaria-specific hospitalised mortality rates declined from 0.43 per 1,000 children p.a. (95% CI: 0.38-0.49) in the 1990s to 0.03 (95% CI: 0.02-0.05; p<0.001) during the period 2020-2024.

**Conclusions:** Hospital admission with malaria among children in Kilifi is now uncommon. Sustained reductions in parasite exposure have altered the clinical profile of disease presentation, including both phenotype and age distribution, without resulting in a cumulative increase in disease burden. Expanded and sustained coverage of effective long-lasting insecticidal nets, together with improved access to effective treatment, have likely contributed to this epidemiological transition.

## Background

The Roll Back Malaria (RBM) initiative was launched in 1998 amid the alarming rise in malaria cases globally [1]. The African Heads of State signed a declaration in April 2000 committing their governments to reverse the malaria trend [2]. A new financing mechanism, the Global Fund to fight AIDS, TB and Malaria, was established in 2002 to increase and consolidate overseas development assistance (ODA) to tackle these three major causes of death, with a primary focus on Africa. This led to unprecedented, transformative interest-free financing for malaria control for many African countries.

Despite billions of dollars in ODA invested in malaria control, most Africa countries have limited direct empirical evidence of changes in disease burden. Infrequent national household surveys are not designed to measure disease incidence or provide reliable sub-national estimates of infection prevalence. Civil registration systems remain weak, and malaria specific mortality is difficult to measure in the absence of pre-mortem parasitological and clinical information [3]. As a result, the global community has relied on combinations of spatial and mechanistic models to predict changes in infection prevalence, morbidity and mortality, often built on sparse data and multiple assumptions [4]. However, models cannot substitute local surveillance and contextual information [5].

Hospital admissions provide a valuable source for empirical disease burden estimation [6]. Severe disease events presenting to emergency care facilities serve as a proxy for life-threatening events in the community that could be averted through timely treatment and effective interventions. Long-term hospital-based data of severe malaria is rare in Africa, with only two sites maintaining continuous surveillance since the launch of RBM, Manhica (Mozambique) [7] and Kilifi (Kenya) [8–10].

This study presents a comprehensive analysis of 35 years of paediatric malaria admissions in a rural population in Kilifi County, representing the longest continuous surveillance of hospitalised malaria in Africa. Temporal changes in disease burden and clinical epidemiology are examined in the context of evolving antimalarial treatment and vector control strategies, providing a perspective on how intervention strategies have shaped changing patterns of malaria risk and insights for future policy.

## Methods

### Study site

The study area is located along the Kenyan coast, within Kilifi County (Additional file 1). The KEMRI-Wellcome Trust Research programme was established in 1989, initiating a series of studies, including longitudinal clinical surveillance of severe malaria at Kilifi County Hospital (KCH) and a community-based randomised controlled trial of insecticide-treated bed net (ITN) between 1993 and 1995 [11]. In April 2002, demographic surveillance was established covering an area of 891 km^2^, extending 39 km north and 32 km south of Kilifi creek (Additional file 1). This area is referred to as the Kilifi Health and Demographic Surveillance System (KHDSS) [12]. The population has been re-enumerated on a rolling basis with rounds every 4 months to capture migration, pregnancies and vital events, alongside re-mapping of newly established homesteads [12]. The urban area immediately surrounding the hospital was excluded because of a high proportion of transient populations from other regions of Kenya (Additional file 1).

### Paediatric ward surveillance

In May 1989, the inpatient surveillance system was established using a standard paediatric admission record form that was completed by physicians to record age, gender, address, illness history, clinical investigations and laboratory results.

Surveillance has been conducted continuously 24/7 for all admissions to the paediatric ward and high-dependency unit. Malaria microscopy was performed on all admissions using 3% Giemsa-stained thick and thin blood films and parasite densities were computed based on the red or white blood cell counts. Full blood counts, including haemoglobin, were obtained using automated haematological analysers supplied by Beckman Coulter. Level of consciousness was assessed using the Blantyre Coma Score (BCS). Final diagnoses were assigned at discharge or death by a team of physicians using ICD-9, and later ICD-10 classifications, following review of all clinical and laboratory results including X-rays, blood and cerebrospinal fluid cultures, haematology and treatment responses. Up to two diagnoses were recorded and, when both completed, they were treated as primary and secondary or co-primary, co-morbid conditions. Clinical and treatment guidelines for paediatric admissions have been consistent with changes in national and WHO policy recommendations. For the present analysis, all admissions aged 1 month to 14 years of age who were residents of the rural KHDSS were included from January 1990 to December 2024.

### Outcomes

Malaria admissions were defined as hospitalisations with a positive malaria blood slide and a discharge diagnosis of malaria recorded as primary, secondary, or co-morbid.

Among malaria admissions, severe malaria phenotypes with complete data since 1990 were defined as follows: a) severe malaria anaemia (SMA), haemoglobin of < 5gm/dl; b) cerebral malaria (CM), BCS < 3; c) hyper-parasitaemia, parasite density >250,000/μl; and d) death during admission. These phenotypes may overlap, and analyses were based on events rather than individual children. Other WHO defined severe malaria manifestation [13], including respiratory distress, hypoglycaemia, prostration, multiple seizures and acute kidney injury were not systematically documented over the surveillance period and were therefore excluded.

### Statistical analysis

#### Community parasite prevalence

Community malaria exposure within the rural KHDSS was estimated from infection prevalence among children admitted for trauma (road traffic accidents, fractures, burns, accidental poisoning with insecticides or kerosene), elective surgery (e.g. circumcision repair, rectal prolapse, dental and hernia repair), snake and animal bites, and neoplasms [8–10]. Annual parasite prevalence was age-standardised to the 2-10 year age group [14], with confidence intervals estimated using 1,000 bootstrap iterations.

#### Annualised hospital admission incidence

Population denominators for children aged 0-14 years were obtained from rural KHDSS census enumerations conducted between 2003 and 2024. Populations for 1990-2002 were back-calculated using an annual population growth rate of 2.97% [12]. For years affected by health workers strikes, 2016-17 and 2021, population denominators were adjusted to account for strike duration. The age structure of annual populations was estimated using observed age proportions from combined census data since 2003, with the first year of life adjusted by 11/12.

Incidence was defined as minimum community-acquired hospital events per 1000 resident children per annum (p.a.), acknowledging that some events may not reach hospital. Annual incidence rates of malaria admissions were estimated using Poisson regression model, with malaria case counts as the dependent variable, year as the independent variable and population as an offset. Similar models were fitted for severe malaria phenotypes (CM, SMA, hyper-parasitaemia and malaria related deaths).

#### Context milestone periods

Since 1990, Kilifi has undergone multiple changes in malaria treatment, vector bionomics and vector control policies, these contextual factors are described in detail in Additional file 2 and five significant context periods are summarised in Table 1.

**Table 1:** Five major periods defined based on insecticide treated nets/long-lasting insecticidal nets (ITN/LLIN), vector dynamics, drug resistance, case-management policy and rainfall since 1990 (see Additional Files 2 and 3 for detailed context descriptions).

| Period | Major threats and intervention milestones |
| --- | --- |
| **1990-1996** | Failing chloroquine (CQ) available through the retail sector and widely used presumptively to treat fevers; gradual ITN introduction as part of a trial in the northern KHDSS; vectors were predominantly *An.gambiae* s.s. biting indoors |
| **1997-2005** | CQ replaced by Sulphadoxine-Pyrimethamine (SP) for first line treatment, available in the retail sector for presumptive treatment, but emerging rapid treatment failures across this period; ITN distributions and net re-treatments using social marketing resulting in low coverage; behavioral and species changes of dominant vectors favoring *An.funestus* s.l. and *An.arabiensis*; dramatic climate event (El Niño, 1997) resulting in heavy flooding |
| **2006-2011** | SP abandoned, and artemether-lumefantrine (AL) policy introduced only to formal health sector with a complicated testing policy not fully implemented; the first mass distribution LLINs in 2006 and a policy change to provide free routine LLIN access through maternal and child health/antenatal care clinics leading to increased coverage; continued dominant vector composition changes, outdoor/early biting and a low human blood index; relatively low rainfall/droughts (except 2006) |
| **2012-2019** | Expanded Test & Treat (TT) policy with increased distributions of malaria rapid diagnostic tests; three mass LLIN campaign distributions (2012, 2015, 2017) with sustained high LLIN coverage and pyrethroid sensitivity; unusually heavy rain 2019 |
| **2020-2024** | Evidence of high AL and TT health worker treatment policy compliance; initiation of community case management of malaria closer to homes; two mass LLIN campaigns (2021, 2024) with sustained high LLIN use |

Prevalence and disease incidence have been assessed across these contextual periods using a Poisson regression model with 1990-1996 as the reference. Changes in median age distributions of malaria admissions and phenotypes were evaluated using the Kruskal-Wallis rank-sum test. Malaria case fatality rate (CFR) was calculated as the proportion of deaths among malaria patients who neither self-discharged nor were transferred to other health facilities. Differences in CFR across the five contextual periods were assessed using a binomial regression model, with 1990-1996 as the reference. All statistical analyses were conducted with R (version 4.4.2).

## Results

Between January 1990 and December 2024, 141,384 paediatric admissions were observed. Exclusions included 25,121 (17.8%) neonates, nine (0.01%) children aged >14 years, 218 (0.2%) with missing age, 3,553 (2.5%) with missing address, and 43,318 (31%) who were not resident of rural KHDSS. Of the 69,165 admissions included in the analysis, 2,654 (3.8%) were missing blood slide results.

### Community parasite prevalence

There were 4,443 (6.7%) admissions aged 1 month-14 years from the rural KHDSS that were admitted for trauma, elective surgery, bites and neoplasms and had a blood slide taken. These data were used to estimate the annual age-standardised parasite prevalence, representing community malaria transmission intensity. Prevalence increased between 1990 and 1992, then remained stable at 35-45% through to 1999.

From 2000 onwards, parasite prevalence declined steadily reaching an average of 5% (95% CI: 3, 6; p<0.001) between 2006 and 2011. It remained below 10% thereafter, with a modest resurgence observed between 2012 and 2015. In the most recent five-year period (2020-2024), community prevalence averaged 2% (95% CI: 1, 4; p<0.001) indicating a further reduction in exposure to infection among children across the rural KHDSS (Figure 1A; Table 2).

**Figure 1:**
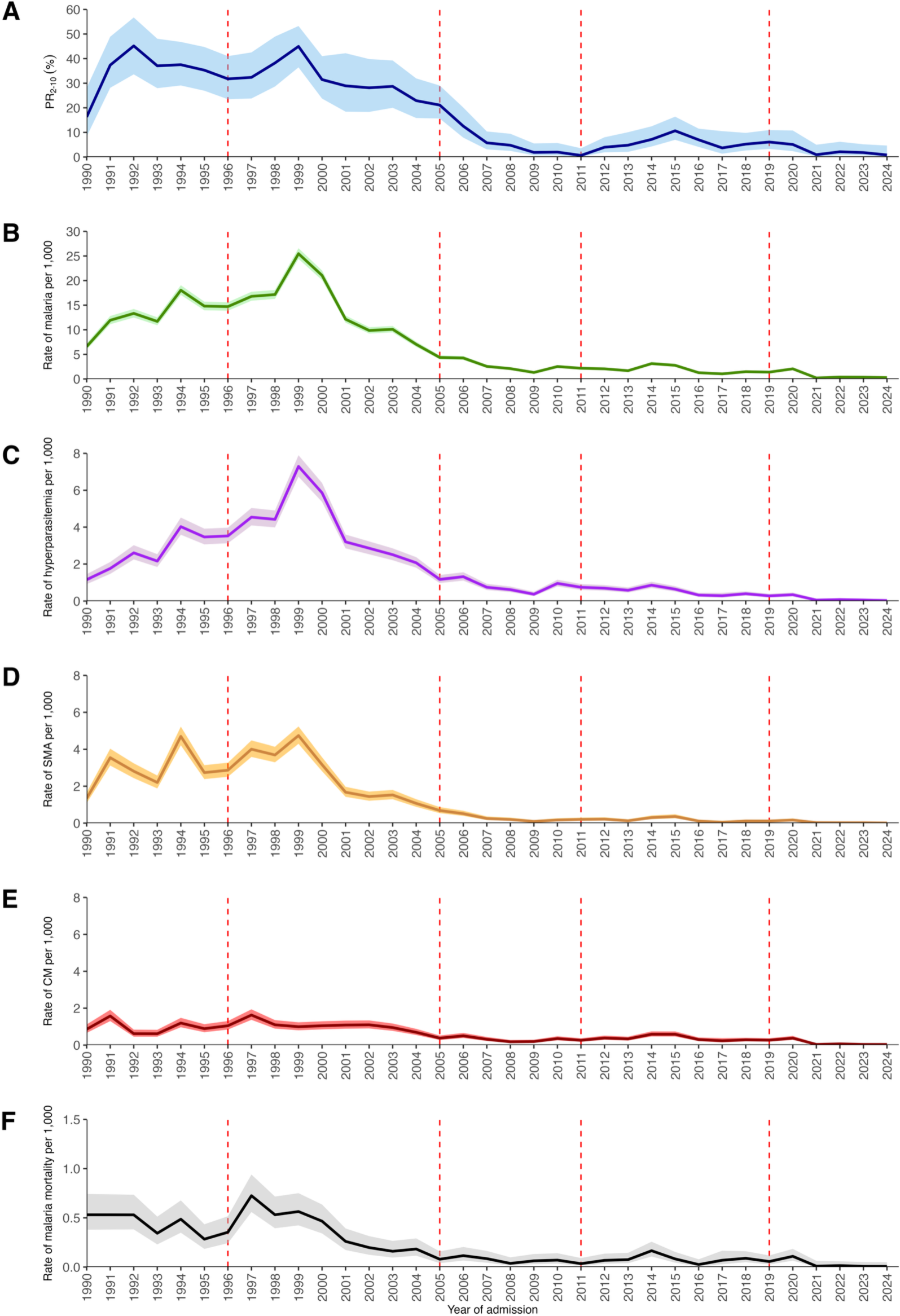
Changing patterns 1990-2024 in background parasite prevalence (A), admission rates per 1000 children p.a. of all-cause malaria (B), and malaria phenotypes (hyper-parasitaemia (C), severe malaria anaemia (D), cerebral malaria (E) and in-hospital mortality due to malaria (F). Dashed red horizontal lines show the five-significant context periods described in Table 1. **Footnote**: Among malaria admissions, parasite density was missing in 73 (0.3%) cases, haemoglobin in 331 (1.6%) and an assessment of consciousness was missing in 193 (0.9%) cases.

**Table 2:** Summary statistics on background parasite prevalence, all-cause malaria, malaria phenotypes (hyper-parasitaemia, severe malaria anaemia (SMA), and cerebral malaria (CM)) and malaria specific mortality across five significant context periods described in Table 1.

|  | 1990-1996 | 1997-2005 | 2006-2011 | 2012-2019 | 2020-2024 |
| --- | --- | --- | --- | --- | --- |
| Trauma/surgical admission age-corrected (2-10 years) SPR; (CI), [N]; p-value‡ | 35%<br>(31, 39)<br>[646] | 34%<br>(30, 37)<br>[925];<br>p=0.14 | 5%<br>(3, 6)<br>[951];<br>p<0.001 | 7%<br>(5, 9)<br>[1,237];<br>p<0.001 | 2%<br>(1, 4)<br>[684];<br>p<0.001 |
| Acute admissions with a diagnosis of malaria at discharge, % (CI) (n/N) | 43.9%<br>(43.2, 44.8)<br>[6,452/<br>14,679] | 41.7%<br>(41.1, 42.3)<br>[10,686/<br>25,604] | 15.7%<br>(15.0, 16.4)<br>[1,623/<br>10,323] | 20.6%<br>(19.7, 21.5)<br>[1,735/<br>8,435] | 8.1%<br>(7.4, 8.8)<br>[432/<br>5,357] |
| Malaria admission rate per 1000 p.a. (CI), [N]; p-value¶ | 13.2‰<br>(12.8, 13.5)<br>[6,452] | 13.3‰<br>(13.1, 13.6)<br>[10,686];<br>p=0.37 | 2.47‰<br>(2.35, 2.59)<br>[1,623];<br>p<0.001 | 1.89‰<br>(1.80, 1.98)<br>[1,735];<br>p<0.001 | 0.65‰<br>(0.59, 0.72)<br>[432];<br>p<0.001 |
| Median age in months (IQR) of malaria admissions [N] | 19 (9, 35)<br>[6,452] | 23 (12, 39)<br>[10,686] | 35 (23, 54)<br>[1,623] | 48 (28, 78)<br>[1,735] | 43 (26, 77)<br>[432] |
| Hyper-parasitaemia malaria admission rate per 1000 p.a., (CI), [N]; p-value¶ | 2.72‰<br>(2.58, 2.87)<br>[1,334] | 3.65‰<br>(3.52, 3.79)<br>[2,925];<br>p<0.001 | 0.79‰<br>(0.72, 0.86)<br>[517];<br>p<0.001 | 0.52‰<br>(0.48, 0.57)<br>[477];<br>p<0.001 | 0.11‰<br>(0.09, 0.14)<br>[73]; p<0.001 |
| Median age in months (IQR) of hyper-parasitaemia admissions [N] | 23 (11, 39)<br>[1,334] | 26 (14, 42)<br>[2,925] | 36 (25, 54)<br>[517] | 49 (31, 80)<br>[477] | 44 (29,69)<br>[73] |
| SMA admission Rate per 1000 p.a. (CI) [N]; p-value¶ | 2.91‰<br>(2.76, 3.06)<br>[1,425] | 2.34‰<br>(2.24, 2.45)<br>[1,877];<br>p<0.001 | 0.23‰<br>(0.20, 0.27)<br>[152];<br>p<0.001 | 0.18‰<br>(0.15, 0.21)<br>[163];<br>p<0.001 | 0.05‰<br>(0.03, 0.07)<br>[31];<br>p<0.001 |
| Median age in months (IQR) of SMA admissions [N] | 15 (7, 32)<br>[1,425] | 17 (9, 31)<br>[1,877] | 28 (19, 42)<br>[152] | 34 (18, 58)<br>[163] | 35 (23, 50)<br>[31] |
| CM admission Rate per 1000 p.a. (CI), [N]; p-value¶ | 0.97‰<br>(0.89, 1.06)<br>[476] | 0.98‰<br>(0.91, 1.05)<br>[781];<br>p=0.94 | 0.30‰<br>(0.26, 0.34)<br>[195];<br>p<0.001 | 0.38‰<br>(0.34, 0.42)<br>[347];<br>p<0.001 | 0.11‰<br>(0.09, 0.14)<br>[71];<br>p<0.001 |
| Median age in months (IQR) of CM admissions [N] | 27 (15, 45)<br>[476] | 27 (15, 40)<br>[781] | 38 (27, 58)<br>[195] | 57 (35, 82)<br>[347] | 52 (32, 94)<br>[71] |
| Ratio CM/SMA | 0.33 | 0.42 | 1.30 | 2.17 | 2.20 |
| In-patient malaria case-fatality rate (CRF), % (CI) (n/N); p-value‡ | 3.29%<br>(2.88, 3.75)<br>[212/6,452] | 2.52%<br>(2.24, 2.84)<br>[269/10,668];<br>p=0.003 | 2.72%<br>(2.03, 3.63)<br>[44/1,619];<br>p=0.24 | 4.25%<br>(3.39, 5.32)<br>[73/1,717];<br>p=0.05 | 4.41%<br>(2.83, 6.81)<br>[19/431];<br>p=0.21 |
| Malaria mortality rate per 1000 p.a. (CI), [N]; p-value¶ | 0.43‰<br>(0.38, 0.49)<br>[212] | 0.34‰<br>(0.30, 0.38)<br>[269];<br>p=0.006 | 0.07‰<br>(0.05, 0.09)<br>[44];<br>p<0.001 | 0.08‰<br>(0.06, 0.10)<br>[73];<br>p<0.001 | 0.03‰<br>(0.02, 0.05)<br>[19];<br>p<0.001 |
| Median age in months (IQR) of malaria mortality [N] | 21 (9, 36)<br>[212] | 24 (9, 38)<br>[269] | 36 (24, 51)<br>[44] | 53 (33, 77)<br>[73] | 42 (26, 94)<br>[19] |

‡ The differences between proportions across the five major context periods were assessed using a binomial regression with the 1990-1996 as the reference period
¶ The differences between incidence rate across the five major context periods were assessed using a Poisson regression model, with the population estimates included as an offset and 1990-1996 as the reference period

### Hospitalised malaria

During the 35-year surveillance period, there were 20,928 admissions with a malaria diagnosis, accounting for 34% of admissions; 17,755 (85%) had malaria as the sole diagnosis, 1,950 (9%) had malaria as the primary diagnosis with a secondary condition, and in 1,223 (6%) malaria was a secondary diagnosis. The annual rate of malaria hospitalisation among children aged 1 month-14 years increased sharply during the 1990s. In 1990, the malaria admission rate was 6.6 per 1,000 children p.a. (95% CI: 6.0, 7.2) and malaria accounted for 27% of admissions. By 1999, malaria hospitalisation rates had risen to a peak of 25.5 per 1,000 children p.a. (95% CI: 24.4, 26.6), representing 53% of admissions (Figure 1B). From 2000 onwards, rates began to decline steadily: 21.1 per 1,000 children p.a. (95% CI: 20.0, 22.0) in 2000 to 1.32 per 1,000 children p.a. (95% CI: 1.12, 1.55) by 2009 (Figure 2B).

**Figure 2:**
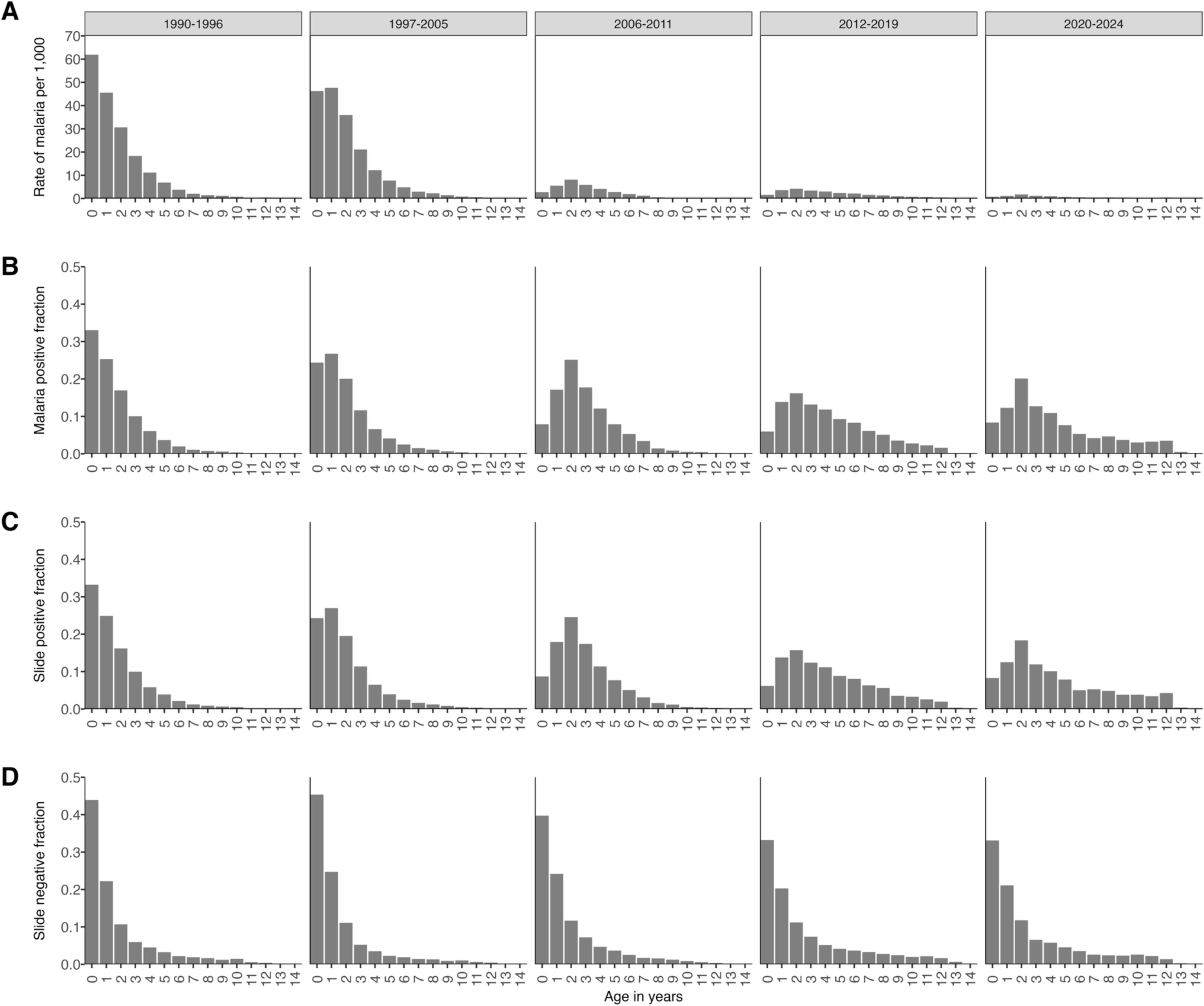
Age patterns of rates of malaria admission (A), fractions of malaria admission (B), slide-positive (C) and slide-negative (D) over five-time intervals among children age 1m-14years at Kilifi County Hospital, 1990–2024.

Across the period of changing intervention (Table 1), malaria accounted for 43.9% (95% CI: 43.2, 44.8) of paediatric admissions in1990-1996, and 41.7% (95% CI: 41.1, 42.3) in 1997-2005. Between 2012 and 2019, malaria hospitalisation rates averaged 1.89 per 1,000 children p.a. (95% CI: 1.80, 1.98; p<0.001) representing only 20.6% of all admissions. The period from 2020 was characterised by a further decline in malaria hospitalisation with an average across the period of 0.65 per 1,000 children p.a. (95% CI: 0.59, 0.72; p<0.001), with only 23-53 malaria admissions between 2021 and 2024 accounting for less than 5% of all admissions (Figure 1B; Table 2).

### Severe malaria phenotypes

Trends in the rates of annual hospitalised malaria admissions with hyper-parasitaemia, severe malaria anaemia and cerebral malaria are shown in Figures 1C-1E. Broadly, the rise and fall in rates of hyper-parasitaemia and SMA were comparable to those described for malaria admission rates (Figure 1B). For example, SMA admission rates at the peak of malaria case burdens in 1999 was 4.74 per 1,000 children p.a. (95% CI: 4.29, 5.23), compared to the lowest admission period, 2020-2024, where the average SMA hospitalisation rate was 0.05 per 1,000 children p.a. (95% CI: 0.03, 0.07; p<0.001) (Figure 1D; Table 2). Although the absolute incidence of cerebral malaria declined over time, the ratio of CM to SMA increased over time, from 0.3 before 2006, increased to 1.3 in 2006-2011 and increased further to 2.2 between 2012 and 2024 (Table 2).

### Hospitalised malaria mortality

Among children admitted with a malaria diagnosis, 617 (2.9%) died before discharge. CFR varied modestly over time, from 3.3% (95% CI: 2.9, 3.8) in 1990-1996 to 2.5% (95% CI: 2.2, 2.8; p=0.003) in 1997-2005. During the lowest transmission periods, the CFR was 2.7% (95% CI: 2.0, 3.6; p=0.24) in 2006-2011 and 4.4% (95% CI: 2.8, 6.8; p=0.21) in 2020-2024. The malaria-specific hospitalised mortality rate was 0.43 per 1,000 children p.a. (95% CI:0.38, 0.49) during 1990-1996 and declined steadily thereafter, reaching 0.03 per 1,000 p.a. (95% CI: 0.02, 0.05; p<0.001) by 2020-2024 (Table 2; Figure 1F).

### Changing age patterns

As transmission intensity and rates of malaria hospitalisation declined, the median age of malaria hospitalisation increased (Figures 2A and 2B; Table 2), rising from 19 months (IQR: 12, 39) between 1990-1996 to 48 months (IQR: 28, 78) between 2012-2019.

Similar changes to the age-specific rates were observed across the three severe malaria phenotypes, with the median age increasing as transmission intensity declined, although the absolute age varied by phenotype (Table 2). The age distribution of malaria shifted over time, with older children increasingly represented in the rate of malaria hospitalisation (Figure 2A) and proportional malaria admissions (Figure 2B), however this shift did not result in an overall increase in rate of malaria hospitalisation (Figure 1B). The age pattern observed for children with a discharge diagnosis of malaria (Figure 2B) was similar to those who were reported as slide positive on admission, regardless of diagnosis (Figure 2C) but slide negative admissions showed a more stable age pattern across all time periods (Figure 2D).

## Discussion

This analysis characterises the long-term trends in the risk and age patterns of severe hospitalised malaria in a rural population on the Kenyan coast over a 35-year period. There has been a 94% reduction in community malaria prevalence since the 1990s (Figure 1A). Consequently, children born within the KHDSS today face a considerably lower risk of malaria infection compared to those born during the 1990s, accompanied by a dramatic decline in malaria hospitalisation from the same community (Figure 1B). Since 1990, the annual burden of hospitalised paediatric malaria at KCH has been fallen from thousands of cases to fewer than 100 per year. Malaria accounted for 43% of paediatric admissions 1990-2005, and since 2021 represents less than 5%. Severe malaria anaemia and hyper-parasitaemia admission rates followed a similar temporal pattern (Figures 1C-1D). Although cerebral malaria now represents a larger proportion of severe cases, the absolute number of cases remain very low (Table 2). In-hospital mortality among children from the KHDSS has declined by 93% since 1990, from 0.43 per 1,000 p.a. (1990-1996) to 0.03 per 1,000 p.a. (2020-2024), and by 63% between 2012-2019 and 2020-2024 (Table 2; Figure 1F).

As transmission intensity and hospitalisation rates declined, the median age of malaria admissions and all slide-positive admissions increased, while the age pattern of children admitted without malaria infection remained constant over the surveillance period (Figure 2). It has been suggested that declining parasite exposure may shift the peak age of severe disease to older children, potentially offsetting overall reductions in risk during childhood [15,16], and that increased vulnerability of older children to cerebral malaria may offset reductions in mortality. This pattern was not observed in the present analysis. As transmission intensity declined, older children had a relatively higher risk of hospitalisation, but the absolute numbers remained low, and this increase did not result in rise or stagnation in cumulative risks (Figure 1; Figure 2; Table 2).

### Plausible contributions to the changing malaria epidemiology

The drivers of changing parasite exposure and disease progression within the KHDSS over the past 35 years are multi-layered and complex (Additional file 2). There are several plausible congruences between policy implementation and changes in infection prevalence and disease incidence. In the late 1990s, two coinciding factors may have accelerated the changing transmission intensity. First, the widespread and easily accessible use of long-half-life, “prophylactic” treatments for fever, sulphadoxine-pyrimethamine (SP). Second, a rapid change in vector composition and behaviour associated with increasing ITN use. This period saw a transition from the historically dominant, anthropophilic *Anopheles gambiae* s.s to more opportunistic vectors (*An. arabiensis* and *An. funestus,* accompanied by increased outdoor biting and a change from almost universal human blood meals in the 1990s to <15% by 2010 [17] (Additional file 2 section 2.3).

Pyrethroid sensitivity bioassay conducted between 1994 and 2014 continued to show high vector mortality to pyrethroids (Additional file 2 section 2.4). Importantly, studies within the KHDSS between 2008 and 2019 consistently showed that long-lasting insecticide treated nets (LLIN) use among children was associated with a 55% reduction in community-based infection prevalence (adjusted OR=0.45; 95% CI: 0.36 to 0.57) [18]; a 32% reduction in out-patient malaria slide positivity (crude OR=0.68; 95% CI: 0.64, 0.73) [19]; and a 33% reduction in malaria admission (HR=0.67; 95% CI: 0.52, 0.85) [20].

Treatment of uncomplicated malaria with chloroquine until 1996 may have contributed to rising hospitalisations, and its replacement with SP, a drug with already reduced efficacy (Additional file 2 section 2.5), did not change the rise in malaria admissions (1997-2005). During the early phases of policy change to artemether/lumefantrine (AL) (2006-2011), treatment access was largely restricted to formal health care providers and adherence to guidelines was sub-optimal (Additional file 2 section 2.6). Following the decentralisation of health services in 2010, there was an increase in health facilities within the KHDSS, alongside renewed efforts to improve malaria case-management improving use and compliance with diagnostics (rapid diagnostic tests (RDT)) and treatment (AL) (2012-2019) (Additional file 2 section 2.6). Since 2023, there has been an expansion of community case management with community health promoters provided with RDTs and AL across the KHDSS (Additional file 2 section 2.7). Evidence from the KHDSS indicates that AL remains an effective first line treatment [21]. Although, the direct effects of prompt, effective treatment on disease progression requiring hospital care are difficult to quantify, it is likely that improved access to effective treatment has contributed to the continued decline in malaria admissions.

Major precipitation anomalies in 1997 associated with El Niño (Additional file 3) coincided with low ITN coverage (Additional file 2 section 2.2) and drug resistance potentially creating a “perfect storm” for malaria transmission. Mean day-time temperatures within the KHDSS have risen by nearly 1°C since 2003 [22]. However, heavy rains events in 2006, 2019 and 2023 (Additional file 3) and rising temperatures since 2003 were not associated with major increases in malaria hospitalisation. While climate change may affect malaria risk at continental scales, within the KHDSS, climate anomalies appear less influential than sustained coverage and effectiveness of long-lasting insecticide treated nets (LLIN), and improved access to effective first-line treatment.

The question therefore is whether the local patterns of malaria since 2006 represent a “new normal”, with infection prevalence below *circa* 5% and malaria hospitalisation rates <5 per 1000 children p.a. In the absence of a sterilising or transmission blocking vaccine, it is unlikely that parasite transmission will be interrupted within the KHDSS in the foreseeable future. A more realistic goal is to prevent malaria deaths. Despite emerging threats from parasite and vector resistance (Additional file 2 section 2.4 and 2.5), this can be achieved through expanded and prompt access to effective treatment, combined with a sustained supply of LLIN. Any adjustments to diagnostics, treatment and vector control must be guided by continuous surveillance of resistance genes, efficacy testing and epidemiological data. Without surveillance at local levels, policy adaptions will lack empirical evidence and are likely to be delayed.

### Conclusion

There has been a sustained, dramatic reduction in paediatric malaria hospitalisation over the past 35 years. During the 1990s, malaria accounted for more than 40% of all paediatric admissions, whereas today malaria requiring emergency care is relatively rare. As hospitalisation incidence has declined, the clinical epidemiology has shifted: children admitted now are older and more likely to have cerebral malaria than severe malaria anaemia. Nevertheless, the overall incidence of hospitalisation and cerebral malaria in older children continues to decline. Despite concerns that progress might stagnate or that disease burden could rise with declining ODA support and emerging resistance threats, this has not been observed in the Kilifi study area. Two core interventions, LLIN and prompt access to AL, have likely been pivotal in reducing severe malaria and associated deaths in this setting.

## List of abbreviations

AL: artemether-lumefantrine; BCS: Blantyre coma score; CFR: case fatality rate; CI: confidence interval; CM: cerebral malaria; HR: hazard ratio; ICD: international classification of diseases; IQR: interquartile range; ITN: insecticide-treated bed net; KCH: Kilifi county hospital; KEMRI: Kenya medical research institute; KHDSS: Kilifi health and demographic surveillance system; KWTRP: KEMRI–Wellcome trust research programme; LLIN: long-lasting insecticide treated nets; MoH: ministry of health; ODA: overseas development assistance; OR: odds ratio; RBM: roll back malaria; RDT: rapid diagnostic test; SMA: severe malaria anaemia; SP: sulphadoxine-pyrimethamine; WHO: World Health Organization

## Ethical approval and consent to participate

This study analysed routinely collected hospital surveillance data. Written informed consent for data collection was obtained at the point of care from parents or legal guardians of children, in line with hospital surveillance procedures. Data used for this analysis were de-identified prior to access by the study team. The hospital surveillance and KHDSS studies were approved by the Kenya Medical Research Institute Scientific Ethics Review Unit (KEMRI/SERU/SSC/1433, KEMRI/SERU/SSC/3057 and KEMRI/SERU/SSC/3149).

## Clinical Trials

Clinical trial number: not applicable.

## Consent for publication

Not applicable.

## Availability of data and materials

These data are available through a formal requesting process to the KEMRI-Wellcome Trust Institutional Data Access/Ethics Committee. Correspondence and requests for materials should be addressed to the KEMRI Wellcome Data Governance Committee.

## Authors’ contribution

RWS and AK conceived the study. NM, SM and GM were responsible for the paediatric ward surveillance. DW and AM led the field activities and data collection within the KHDSS with oversight from TNW. AK led the data analysis with assistance from MMM. AK and RWS interpreted the data and drafted the manuscript, with input from TWN and PB. All authors had full access to all the data in the study and had final responsibility for the decision to submit for publication. All authors read and approved the final version of the manuscript.

## Acknowledgments

We are grateful to all the residents of Kilifi who have participated in the surveillance activities of the KHDSS since 1990. We acknowledge the enormous effort provided by the KWTRP nurses, clinical officers and doctors who have provided cover on the paediatric ward over many years, the census field staff and data supervisors who collect and process the community information and the data entry staff who ensure its constant updating and accessibility, finally we acknowledge the Community Liaison Group who ensure that this work can operate with a true community engagement. We would like to further acknowledge the valuable insights related to important aspects of the context described in this paper (Additional file 2). We wish to thank Dejan Zurovac, Emelda Okiro, Peter Macharia and Isabella Oyier-Ochola for comments on an earlier version of the manuscript.

