## Additional file 1 for "Decline to near-zero malaria hospitalisation over 35 years on the Kenyan Coast"

**Additional File 1: Kilifi study area and population**

Studies on severe malaria and its epidemiology in the area surrounding Kilifi hospital (KCH) began during the period 1989-1993 with Wellcome Trust funding to the University of Oxford in collaboration with the Kenya Medical Research Institute (KEMRI) including descriptions of the clinical spectrum of severe paediatric malaria [1], severe anaemia [2], the validity of Verbal Autopsies in the attribution of paediatric causes of death [3], case-control studies on severe disease progression [4,5] and parallel, allied studies related to entomological risk factors [6] and human behaviour [7]. The research platform enabled linkage of hospitalised events to locations and sub-locations surrounding KCH. Demographic surveillance for the entire study area was established in 2002 [8] enabling linkage to resident household members living in 186 enumeration zones (EZ) across 15 locations (Figure 1). The rural population in 2003 was 183,509 and grew to 286,078 by 2024. Continuous community engagement activities have been maintained for over 20 years to explain the purposes of the research to the community [9,10].

Most residents of the KHDSS belong to the Mijikenda group while other ethnicities have settled in the area from Central and Western Kenya with a notable mixture of different groups in Kilifi Township. Among the rural population, agriculture forms the basis of most household activity. Adult male household members who work outside the KHDSS provide an important source of financial remittance for the community. Kilifi Township, and the area immediately south of the creek connected by a bridge (Mnarani), has grown significantly since 1990 and comprise traders, artisans, county government and public service administrators, police, hospital personnel and those employed in the transport and tourist industries. Kilifi township includes a hospital maintenance school that provides training for students from across Kenya, the KEMRI research campus grew significantly from 1990 and now provides employment for over 700 staff, Pwani University was established in 2007 and provides courses for *circa* 8000 students all housed within the township area, there are four international hotels and the Kilifi Sisal Plantation is located south of Kilifi Creek. These centres provide employment for people from across Kenya in the township area. This area (Figure 1; dark grey) is characterised by substantial population mobility and a high proportion of transient residents. Because these dynamics complicate the attribution of hospitalisations to a well-defined, stable catchment population, admissions from this area were excluded.

**Figure 1:** Location of Kilifi Health and Demographic Surveillance Site showing location of Kilifi County Hospital (KCH), the urban extents excluded from analysis (dark grey), households (light grey dots) and the main Malindi-Mombasa road (red). Grey boundaries show administrative locations.

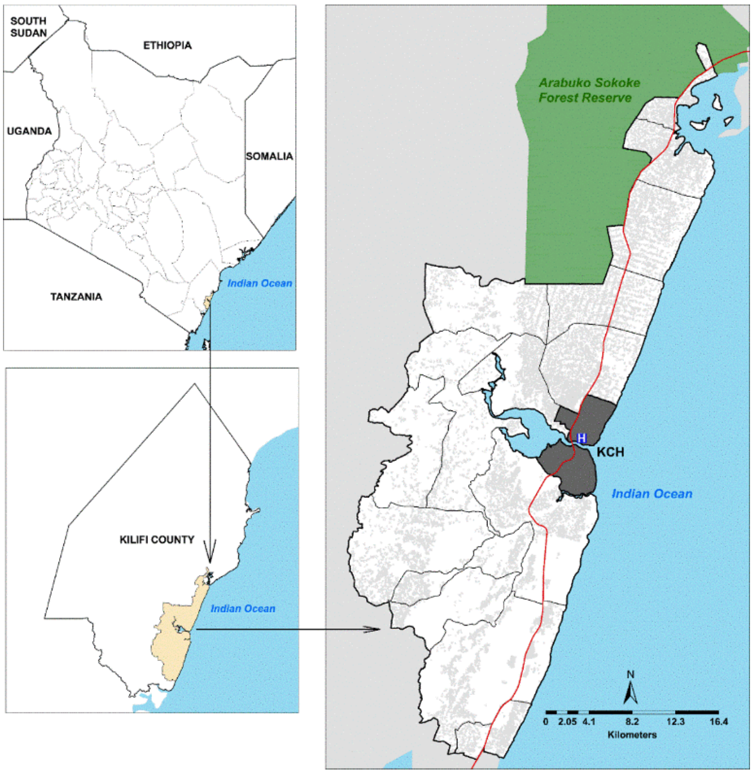
