## Additional file 2 for "Decline to near-zero malaria hospitalisation over 35 years on the Kenyan Coast"

**Additional File 2: Context**

### 2.1. Kenya context

Before the Roll Back Malaria (RBM) initiative was launched in 1998, Kenya relied on World Bank loans and bilateral Overseas Development Assistance (ODA) to support malaria control. During the 1990s, assistance to the Division of Vector Borne Diseases, Ministry of Health (MoH) supported community-based initiatives, limited indoor spraying in epidemic-prone areas, vector surveillance, and drug resistance monitoring. By 2000, bilateral support facilitated the transition from an integrated vector-borne disease programme to a dedicated Division of Malaria Control within the MoH and the development of the first post-RBM national malaria strategy in 2001 [1]. This strategy aligned with RBM principles to expand insecticide-treated bed net (ITN) use and improve case management, aiming to reduce malaria mortality by 30% by 2006 [1].

Since 2004, Kenya has received over USD 1.5 billion in ODA specifically allocated to malaria control [2]. These funds have supported important changes in the delivery of ITN, transitions from monotherapies to artemisinin-based combination therapy, increasing access to diagnostics, coverage of intermittent presumptive treatment of malaria in pregnancy, geographically restricted use of indoor residual house-spraying, and periodic (every 3-5 years) national household surveys to track progress in intervention coverage and infection prevalence. In 2010, Kenya was one of the first African countries to adopt a sub-nationally tailored approach to intervention, based on its diverse malaria ecology, focusing malaria prevention efforts on high burden regions around Lake Victoria and along the Kenyan coast and effective case-management nationwide [3]. National strategic plans have set progressively ambitious targets for reducing malaria mortality by 66% between 2010 and 2015 [3], 75% between 2019 and 2023 [4] and 90% 2023 to 2027 [5], but evaluation of these targets has been less explicit.

### 2.2. Insecticide treated nets within the KDHSS

At the end of the 1980s bed net use in the KHDSS was low (6%) [6]. Between June 1993 and July 1995, coverage rose to about 25% of the current KHDSS during a community randomised controlled trial of permethrin treated nets [7]. Control communities were provided nets and re-treatment services from October 1995 [8]. Net re-treatment was provided using social marketing approaches between 1996 and 1997 and treated net coverage declined back to <7% [8]. In January 2002, the UK Department for International Development (DFID) supported Population Services Internation (PSI)-Kenya to socially market partially subsidised nets and re-retreatment packages through the retail sector. This represented the only source of insecticide treated nets (ITN) in Kenya between 2002 and 2004. Coverage of reported ITN use in 2002-2003 within the KHDSS was estimated to be *circa* 7% [9]. In June 2004, DFID extended support to PSI-Kenya to establish a parallel distribution system of heavily subsidised *Supanets* bundled with *Powertab* net treatment tablets (for every 6 months) and supplied to children and pregnant women through Maternal and Child Health (MCH) clinics; this programme began in Kilifi in October 2004. The products were changed to long-lasting insecticidal nets (LLINs), *Olyset* and *Permanet* (permethrin pre-treated nets) in May 2005. Surveys undertaken in Kilifi District in October 2005 reported 14% ITN use among the population. The dual system of socially marketed nets and subsidised branded nets through MCH continued to be the only source of LLIN in Kenya until 2006. In July 2006, free-mass distributions of LLIN were combined with community distribution points for measles vaccine nationwide catch-up campaigns. From 2007 the Kenyan Government began providing LLIN free of charge to pregnant women and children under five years in highly endemic regions, including Kilifi, through antenatal care (ANC) and MCH clinics. The national Malaria Indicator Survey in August 2007 included eight clusters located in the KHDSS where 37% of household residents reported using an LLIN [10]. Subsequent mass LLIN “catch-up” campaigns in Kilifi County occurred in March 2012, September 2015, November 2017, May 2021 and July 2024 (Figure 1).

From 2008, data has been collected annually as part of household re-enumerations of the KHDSS and reported LLIN use among all age groups was 49-64% between 2008-2012, 56-83% between 2013-2017 and 71-90% between 2018-2024 (Figure 1). All LLIN distributed within the KHDSS since 2006 have been pyrethroid treated nets provided free of charge. Piperonyl butoxide (PBO) nets have not been used in coastal counties and within the KHDSS, indoor residual spraying or larval source management have not been used as a vector control measures since 1990.

**Figure 1**: Evolution of ITN/LLIN delivery approaches (top bar), mass-campaigns shown as red lines and estimate coverage (reported sleeping under a treated net the night before the survey) among all age-groups in the KHDSS

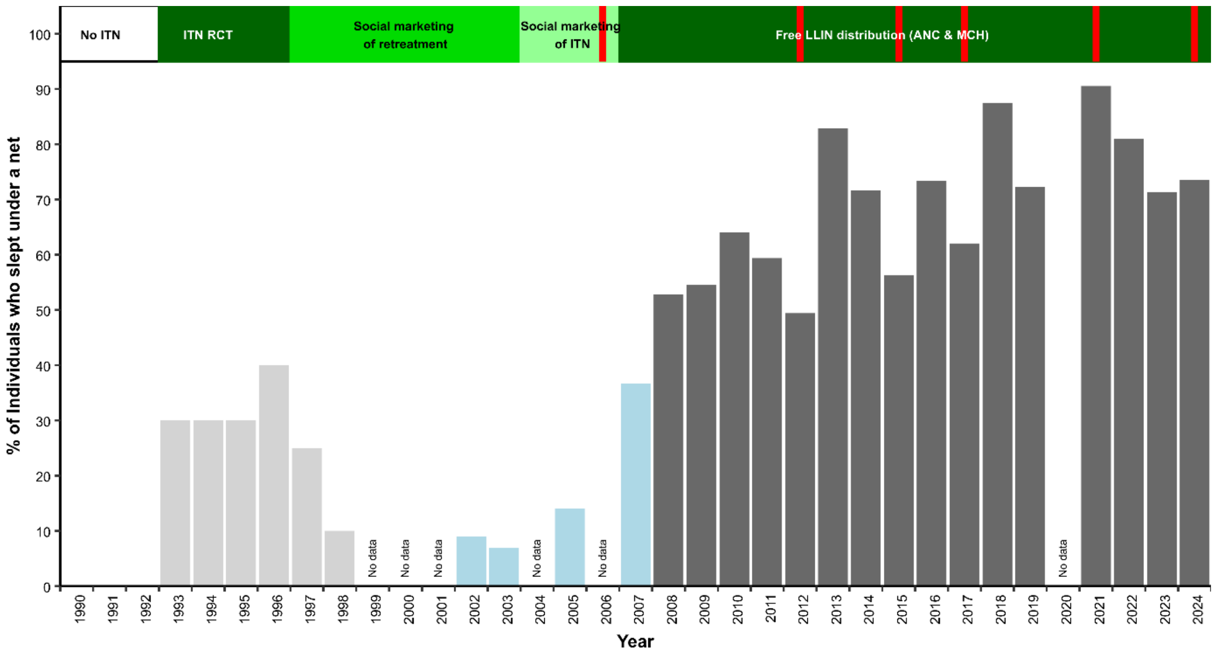


During the 1990s, Kilifi was one of the sites included in a WHO sponsored, multi-site community randomised controlled study to assess the mortality impact of ITN, and the only site to document the impact on severe, hospitalised malaria. Between 1993-1995, the trial demonstrated that infection rates were reduced by 50% [11] and corresponded to a 44% (95% CI: 19-62) protection against the incidence of severe paediatric malaria presenting to KCH [7].

#### 2.3. Vector bionomics

During the 1990s, within the KHDSS dominant vector species sampled indoors included *An. gambiae* s.s. (76%), *An. arabiensis* (10%), *An. merus* (5%) and *An. funestus* s.l. (8.5%) [15]. Other species identified within the KHDSS include *An. pretoriensis*, *An. coustani*, *An. moucheti, An. nili, An. pharoensis* and *An. squamosus* [16–18], whilst implicated as secondary vectors elsewhere they have not been directly identified as playing a role in transmission within the KHDSS. By 1997-1998, following the ITN trials, indoor resting vectors showed a shift toward *An. funestus* s.l. (56%) versus *An. gambiae* s.s. (42%), *An. arabiensis* (2%) and *An. merus* (1%) [19] and a reduced presence of *An. gambiae* s.s (4%) continued though to 2016 [13]. During the ITN trial (1993-1995) there was a significant, immediate behavioural shift among the predominant vector *An. gambiae* s.s toward out-door biting [15]. This phenotypic adaptation continued though to 2016, with over 50% of adult female vectors sampled outdoors and *An. funestus* s.l. biting later in the morning [17]. Between 1990 and 2010, both *An. gambiae* s.l. and *An. funestus* s.l. exhibited a shift from human to animal feeding, declining from 99% to 16% and 100% to 3%, respectively [20]. The efficient, invasive species of *An. stephensi* and *An. gambiae s.s.* M-form (*An. coluzzii*) have not yet been reported within the KHDSS [Martin Rono, personal communication].

**2.4. Insecticide resistance**

In 1994-1995, bioassay results during the ITN trials showed >96% 24-hour post-exposure mortality [15]. In 2011, bioassays of susceptibility to pyrethroids showed greater than 95% mortality after 24 hours among both *An. gambiae* s.l and *An. funestus* s.l. populations (Mbogo CNM, unpublished data). Between 2013-2014, larvae sampled in Kilifi County, including sites in KHDSS, were reared to adults for bioassays and 24hr mortality showed complete sensitivity to pyrethroids, except in one site outside the KHDSS north of Malindi [21]. In 2016, three sites within the KHDSS showed bioassay 24hr mortality rates of 98% for *An. funestus* s.l. to permethrin and deltamethrin, but between 93-96% for *An. gambiae* s.l. [18]. In 2016, the L1014S *kdr* point mutation was detected only in *An. gambiae s.s.* at a low allelic frequency (3.3%), and the 1014F *kdr* mutation was not detected in either *An. gambiae s.s.* or *An. arabiensis* within the KHDSS [21].

More recent studies, December 2024, at Mbogolo and Marana in Malindi along Sabaki river, north of the KHDSS have shown a much reduced 24-hour bioassay sensitivity among *An. arabiensis* populations to pyrethroids (39% mortality), deltamethrin (91%), however piperonyl butoxide (PBO) plus permethrin or deltamethrin showed 100% mortality after 24 hours [Joesph Mwangangi, personal communication]. In 2023-2024, bioassay resistance testing in Kwale, the county south of Kilifi on the Coast, showed *An. arabiensis* and *An. funestus* s.s. were resistant to permethrin (24 hour bioassay mortality, 59% and 57%, respectively) and deltamethrin (mortality 51% and 76%, respectively), but susceptible to PBO (100%); Both *kdr* west and east were detected in *Anopheles arabiensis* (L1014S freq = 0.83%, L1014F freq = 0.63%) [22].

#### 2.5. Parasite resistance and changing malaria treatment recommendations

In 1987, CQ failed to clear over 40% of out-patient infections within 28 days in Kilifi [23]. In 1987, the parasitological clearance by day 7 was 100% among patients treated with SP at the KCH out-patient department [24]. Over the following 10 years escalating CQ treatment failures were documented across Kenya [25].

CQ was formally replaced as a first line treatment with Sulphadoxine-Pyrimethamine (SP) in 1997-1998 (Figure 2) [25]. However, SP resistance rose rapidly within the KHDSS, parasitological failures by day 7 rose from 2% in 1993 to 20% by 2000 [26]. Both CQ and SP remained available in the retail sector for fever management and specific interventions targeting shopkeepers to improve appropriate dosing and advice were undertaken in the KHDSS between 1996-1999 [27].

**Figure 2:** Summary of treatment policy, guidelines and access

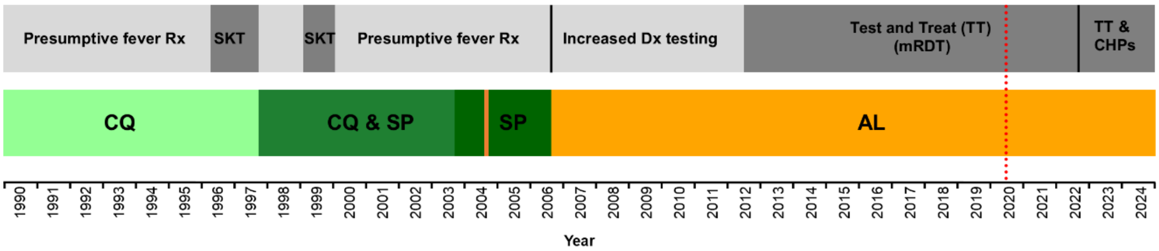


**Footnote**: CQ = chloroquine; SP = sulphadoxine-pyrimethamine, AL = artemether-lumefantrine; SKT = Shop Keeper Training programme; Rx = treatment; Dx = diagnosis; TT = test and treat policy introduced in 2006 and revised in 2010 but more completely implemented in 2012 following extensive roll-out of malaria rapid diagnostic tests (mRDT), prior to 2007 most malaria cases were presumptively diagnosed (Dx) and treated (Rx). The first use of community health promoters to Dx (mRDT) and Rx (AL) as part of community case management of malaria (CCMm) began in 2023. Red dotted line in 2020 represents COVID pandemic.

Data from a co-created network of researchers and ministries of health from the East Africa region mounted sentinel site surveillance of SP effectiveness between 1998 and 2002 [28]. Between 2000 and 2002, day 28 adequate clinical response rates for SP was 85% across at 14 Kenyan sites and 3 sites were below 75% [29]. This evidence was instrumental in supporting the policy change to replace SP with artemisinin-based combination, artemether-lumefantrine (AL) as first-line treatment for uncomplicated malaria in 2004 (Figure 2) [30]. However, the complex procurement, supply, in-service training and raising awareness surrounding a multi-dose regimen led to delays in implementation until 2006 [30].

Drug trials that have compared new comparator drugs with AL within the KHDSS have shown continued efficacy between 2005-2008, day 28 PCR corrected clearance of 99% [31], and between 2018-2019, day 42 PCR corrected clearance of 96% [32]. Genes associated with artemisinin resistance, *Plasmodium falciparum* Kelch 13 (*Pfk13)*, were detected in East Africa in 2015 and have expanded sub-regionally [33]. Emerging *Pfk13* mutations (P553L) were first described at low frequencies (2%) in 2016 in Kisumu. Molecular surveillance on stored samples from KCH admissions for *Kelch13* mutations between 2015-2018, yielded several nonsynonymous mutations, and none of them were validated artemisinin resistance associated mutations [34]. Several *Pfk13* mutations have recently been observed during surveys in 2021 and 2023 of school children within the KHDSS, including two WHO validated artemisinin resistance mutations were identified at low frequencies (<3%), P553L (2021) and S522C (2023) [Victor Osoti personal communication].

Histidine-rich Protein (*hrp*) 2 and 3 gene deletions, allow the parasites to evade detection, resulting in false negative RDT results. Within the KHDSS *Pfhrp2/3* deletions were examined among 345 uncomplicated malaria patients presenting to the Pinglikani dispensary between November 2019 and February 2020 [35]. Blood was tested for infection using mRDT (Carestart), microscopy and qPCR. False negatives were defined in 11 samples that were RDT-negative and microscopy positive and 25 samples that were qPCR-positive and RDT-negative; none had any *Pfhrp2/3* deletions. Separately, 242 qPCR-positive were examined and 5 (2.1%) samples were suggestive of *Pfhrp2/3* deletions [35].

#### 2.6. Treatment guidelines

Prior to 2006, malaria case-management was essentially treatment of all fevers presumptively as malaria across all age groups and levels of care [36,37]. The use of CQ for fever management by communities on the Kenyan coast is likely to have been prolific and spanning several decades before 1990; 70% of cases of malaria sought CQ treatment from retail shops for first line treatment during the early 1990s [38,39]. Given the importance of the retail sector in accessing anti-malarial treatment by communities in the KHDSS, a shopkeeper educational programme was mounted in the Southern part of the KHDSS in 1996-1997 when CQ was the recommended first line treatment [40], and in 1999 in the same area when SP replaced CQ; which expanded to the northern part of the KHDSS at a later implementation phase in 2000 [27]. SP and CQ were withdrawn from the retail sector in 2006, and AL became a prescription-only-medicine only available through formal health facilities [30].

In 2006, when the new AL policy was implemented, the MoH revised recommendations to include parasitological diagnosis at facilities where malaria microscopy or malaria rapid diagnostic tests (mRDT) were available [41], however, all febrile children below 5 years of age in high malaria risk areas were recommended to be presumptively treated with AL and all febrile patients without another obvious cause of fever aged >5 years should have a malaria test performed with treatment for malaria reserved only for only patients who test positive. While testing rates may have increased, adherence to these new guidelines was poor [42]. In 2012, Kenya revised its “test and treat” guidelines [43] to testing all fevers and these guidelines were accompanied by in-service training of all health cadres and provision of new job aides, re-training of microscopists and expanded access to mRDTs through large procurements under the Global Fund awards to Kenya with notable large-scale national distributions of mRDTs from 2012. That same year, child friendly AL dispersible tablets were introduced into the Public Health Sector. The treatment policy was further revised to recommend dihydroartemisinin-piperaquine

(DHA-PPQ) as the second-line treatment. However, DHA-PPQ had not been distributed in the public sector by 2021[44] and by 2024, only 1.8% of out-patient facilities nationwide had DHA-PPQ in stock [45].

Implementation of parenteral artesunate, in the 2012 treatment policy replaced quinine for prereferral and inpatient severe malaria management [43], faced similar stock-outs at peripheral facilities [46]. Health workers' compliance with malaria testing of febrile patients in public facilities improved from 40% (2010) to 76% (2016). By 2016, nearly 100% treatment compliance with test positive and test negative results was observed at facilities in coastal Kenya where AL and malaria diagnostics were available on survey days [47].

#### 2.7. Treatment access

KCH has served as the main level 4 emergency care, in-patient service since 1990 with paediatric bed capacity increasing from 35 and a five-bed high dependency unit in 1995 to over 70 beds on the main ward and 15 beds on the HDU by 2016. All paediatric admissions are managed by KEMRI-Wellcome Programme clinical staff. During the 1990s the standard malaria treatment at KCH for children with evidence of severe malaria was IV quinine (QN). During the 1990s/early 2000s children able to take oral medication received SP plus CQ or pre-discharge following QN treatment. In 2000/2001, oral treatments for uncomplicated malaria admissions were changed from SP/CQ to SP alone. In 2004, IV QN was changed to IV Artemether and oral treatments changed to AL. IV Artesunate became national policy in 2012 for severe malaria [43]. Blood transfusions for severe anaemia (haemoglobin <5gm/dl) have been restricted to children with respiratory distress or children with a 20% infected red cell hyper-parasitaemia and low haemoglobin. In addition, benzyl penicillin and chloramphenicol in patients with neurological impairment until meningitis had been ruled out with microbiology.

Peripheral health facilities (level 2 and 3) managed by the Ministry of Health were few in the early 1990s within the KHDSS (*circa* 9 in 1995) but had increased to 15 by 2006 [48].

In 2021, level 2 and 3 facilities had increased within the KHDSS to 23 [49]. The precise growth in private sector providers is hard to enumerate but have been present since 1990 within the KHDSS providing malaria case-management out-patient services [50].

The Kenya Community Health Strategy 2020-2030 outlines specific roles for Community Health Promoters (CHPs) in diagnosing and treating common childhood illnesses, including malaria [51]. However, implementation of this Level 1 service was slow. In 2023, a formal community case management of malaria (CCMm) programme was launched in Kilifi County, covering six areas and involving 60 CHPs within the KHDSS. The CHPs were trained and provided with supplies for testing (mRDTs) and treatment (AL) to manage children aged 8 months to 5 years (https://www.world-friends.it/en/kenya/).

#### 2.8. Service disruptions and COVID

Since 2010 Kenya has witnessed multiple health worker strikes [52–54]. Significant were strikes in 2016-2017, lasting 100 days (doctors) and 150 days (nurses) and those lasting 70 days in 2021 during the COVID 19 epidemic in response to lack of provision of PPE and health insurance. Within the KHDSS the 2017 strikes impacted on paediatric admission, out-patient visits and ANC attendances between June and October 2017 [53]. However, during strike periods, the HDU at KCH, managed by KEMRI-Wellcome staff, remained operational but admissions to this unit were restricted to the most critical cases. Kenya reported the first case of COVID-19 on 13th March 2020 and “lock down polices” were implemented in Kilifi County between April-June 2020 and curfews were re-started in September 2020 for three months during the second wave of infections and again in March 2021 during a third peak in infections [55,56]. It is unclear how these COVID responses impacted on hospital admission, other than the related strike events. There were nationwide stock outs of mRDTs in 2013 because 4.4 million mRDTs were destroyed by a fire at the Kenya Medical Supplies Agency (KEMSA) stores. No data are available on the impact of this commodity supply interruption within the KHDSS.

#### 2.9. Acknowledgements

We acknowledge several individuals who provided additional detail on the context of control and treatment used here: Guantai Dickson, Patricia Cidi and Chris Wilson (Kilifi Plantation), Andrew Wamari, Dejan Zurovac, Beatrice Amboko, Moses Chapa, Isabella Ochola-Oyier, Victor Osoti, Nancy Kagwanja, Charles Mbogo, Janet Midega, Joesph Mwangangi, Martin Rono, Moriasi Nyanchoka and Benjamin Tsofa.

31. Borrmann S, Sasi P, Mwai L, Bashraheil M, Abdallah A, Muriithi S, et al. Declining responsiveness of Plasmodium falciparum infections to artemisinin-based combination

treatments on the Kenyan coast. PLoS One. 2011;6:e26005. https://doi.org/10.1371/journal.pone.0026005

43. MoPHS. National Guidelines for Diagnosis, Treatment and Prevention of Malaria for Health Workers. Nairobi: Ministry of Health; 2012.
