## Additional file 3 for "Decline to near-zero malaria hospitalisation over 35 years on the Kenyan Coast"

**Additional File 3: Climate**

Kenya's coastal climate is the Inter-tropical Convergence Zone of the Northeast and Southeast Trade Winds. The hot northeast monsoon occurs from November to March/April and includes the 'short rains', variably between November and December. The moist monsoon blowing in from the southeast occurs from April/May to October leading to the heaviest, 'long rains' (March, April, May and June). The rainfall patterns exhibit large inter-annual variability. Monthly rainfall has been recorded continuously at the sisal plantation 2 km south of the creek. Significant rainfall anomalies were noted during the El Niño flooding of 1997 that impacted malaria burdens nationwide. Further periods of high rainfall were recorded during 2006, 2019 and 2023. The period 2008-2016 experienced relatively low rainfall and again during 2021-2024 period, except 2023 (Figure 1). Temperatures range between a minimum of 23°C in April, May and June to a maximum of 30°C in December and January. There has been a gradual warming within the KHDSS with an average of 0.049°C increase per year, and a net temperature increase of 0.98°C over 20 years (2003-2023) as defined by remotely sensed land-surface temperatures from the Moderate Resolution Imaging Spectroradiometer sensor [1].

**Figure 1**: Annual rainfall recorded at the Kilifi Plantation located 2 km south of Kilifi creek. Dotted red line represents the mean annual rainfall over the 35 years of surveillance

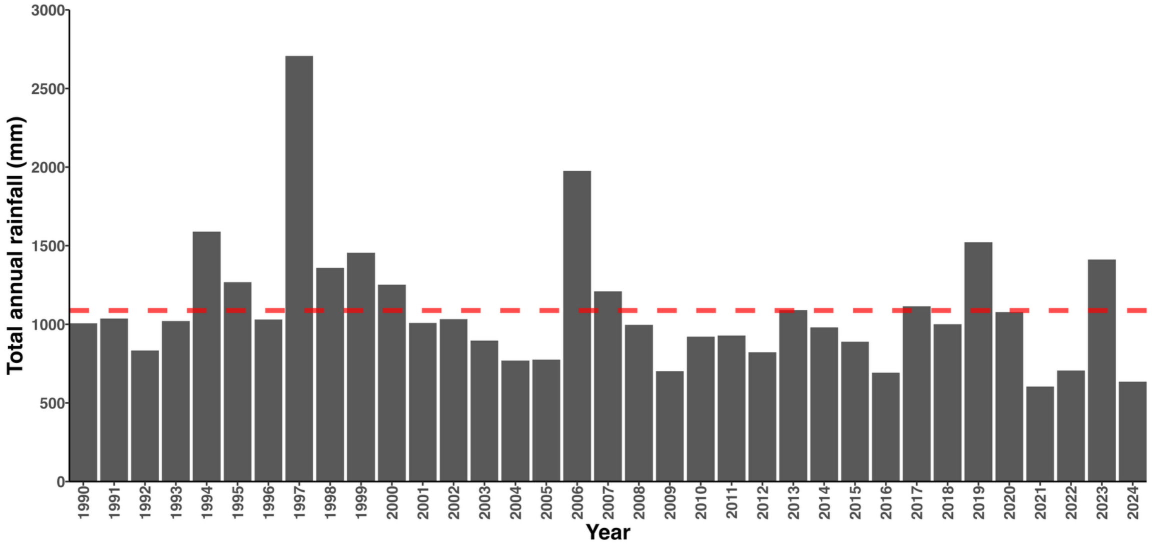
